# Bi-directional associations between mask usage and the associated reasons before and after the downgrading of the legal status of COVID-19 in Japan: A longitudinal study

**DOI:** 10.1101/2023.07.28.23293298

**Authors:** Michio Murakami

## Abstract

**Objectives:** From a public health perspective, it is important to clarify the associations between mask usage and the associated reasons in situations when mask usage is promoted or mitigated. Therefore, I clarified the changes in mask usage and the associated reasons before and after the downgrading of the legal status of COVID-19 in Japan, and analyzed the bi-directional associations between the two.

**Design:** Longitudinal study.

**Methods:** Online surveys were conducted in two waves, between April 18–19, 2023 and June 6–15, 2023, among people aged 20–69 years living in Japan. A total of 291 participants completed both the surveys. The associations between mask usage and beliefs about the reasons for mask usage were analyzed using a cross-lagged panel model.

**Results:** Mask usage decreased slightly, but significantly, from the first to the second wave (*P* < 0.001, Cohen’s d = −0.23). Of the eight beliefs regarding mask usage, slight but significant decreases were observed in terms of relief and information effects (*P* = 0.046, Cohen’s d = −0.12; *P* = 0.018, Cohen’s d = −0.14). There was a significant association between socio-psychological reasons other than infection risk avoidance (such as norm and relief) during the first wave and mask usage during the second wave [standard estimates:0.25 (95% confidence interval (CI):0.06–0.44)]. Contrarily, mask usage during the first wave was significantly associated with the reasons for infection risk avoidance during the second wave [standard estimates:0.13 (0.03–0.24)].

**Conclusions:** The impact of downgrading the legal status of COVID-19 in Japan on mask usage and the associated reasons were found to be limited. In terms of promoting or mitigating mask usage, the significance of risk communication based on socio-psychological reasons other than infection risk avoidance, such as norms and relief, was highlighted.

## INTRODUCTION

In addition to vaccination and social measures such as lockdown, measures against infectious disease outbreaks include infection control at individual level using steps such as hand washing and mask usage, which are less invasive. In the aftermath of coronavirus disease 2019 (COVID-19) outbreak, mask usage was one of the measures emphasized upon. In the early phase of the COVID- 19 outbreak (i.e., in the first half of 2020), mask usage was internationally recommended for prevention of transmission ^1^. Moreover, based on the findings of hydrodynamic studies on airborne droplet particles ^2^ and a meta-analysis of epidemiological studies regarding reduction of the risk of infection by wearing masks ^3^, mask usage has also been contemplated to have the effect of preventing infection for the person wearing it. Furthermore, randomized controlled trials on risk reduction of COVID-19 by mask usage at community level ^4^ and infection risk reduction effect using difference-in-differences method ^5^ have supported the utility of mask usage at population level. Contrastingly, disadvantages of mask usage have also been reported, which include physical effects such as discomfort, breathlessness, and headache ^6^. In addition, psychological effects such as lack of empathy and decreased communication have also been reported ^6^.

On May 5, 2023, the World Health Organization (WHO) declared the end of COVID-19 as a global health emergency ^7^. Even though mask usage was not mandatory in Japan and based on a request from the government, Japan has one of the highest proportion of mask usage in the world ^8^. On March 13, 2023, Japan changed its policy from the previous recommendation of wearing masks indoors to one where individual judgment was the basis for wearing masks ^9^. Furthermore, on May 8, Japan downgraded the legal status of COVID-19 to the same level as seasonal influenza ^9^. This may have resulted in a change in individual mask-usage behavior in Japan.

A systematic review of the factors contributing to infection prevention behaviors, such as mask usage, reported that the infection prevention behavior was influenced by contextual factors (e.g., confidence in science), socioeconomic position (e.g., annual income), and intermediary determinants (e.g., social norms)^10^. Among these, identifying which of the multiple beliefs regarding the reasons for mask usage would be useful in developing risk communication strategies to promote or mitigate mask usage. Nakayachi et al. surveyed Japanese people in March 2020 and reported that among the three reasons related to infection risk avoidance [i.e., perceived severity (severity), perceived self-efficacy of wearing a mask for protection (protection), and perceived efficacy of wearing a mask to prevent spread (prevention)] and three other socio-psychological reasons [perceived norm to wear masks (norm), feeling relief when wearing masks (relief), and impulse to take whatever actions are necessary (impulsion)], norms were most strongly associated with mask usage, followed by relief ^11^. In addition, a randomized controlled trial conducted in Japan in March 2021 also showed that norms were more effective than information provided by medical experts for mask usage ^12^. Contrastingly, a longitudinal study conducted four times in Switzerland between March and July 2020 showed that the belief that masks protect others was strongly associated with mask usage ^13^. Although these studies have investigated the factors during COVID- 19 outbreak when mask usage was recommended, no study has identified the reasons associated with mask usage during periods of decline. Moreover, while these studies analyzed the reasons for mask usage from the perspective of influencing mask usage, it is possible, given a self-perception theory ^14^, that the presence or absence of mask usage may influence beliefs regarding the associated reasons. However, only a limited number of studies have examined the bi-directional associations between mask usage and beliefs regarding the associated reasons. A recent longitudinal study over a 2-year period beginning in the fall of 2020 in the Unites States showed significant bi-directional associations between mask usage and norms ^15^. Therefore, examining the associations between the two entities before and after the downgrading of the legal status of COVID-19 in Japan from a bi- directional perspective would be crucial in developing strategies not only to promote infection prevention measures in another outbreak of COVID-19 or an emergent infectious disease outbreak, but also in mitigating mask usage.

The purpose of this study was twofold. First, I determined the changes in mask usage and beliefs regarding the associated reasons before and after the downgrading of the legal status of COVID-19 in Japan. Second, I analyzed the bi-directional associations between mask usage and beliefs regarding the associated reasons.

## METHODS

### Ethics approval

This study was approved by the Ethics Committee of the Center for Infectious Disease Education and Research, Osaka University (approval number 2022CRER0307). Participant consent was obtained before commencing the survey. This study only included participants who checked the consent box online.

### Study participants

This was a longitudinal study. Two waves of surveys were conducted to monitor individuals aged 20–69 years living in Japan who were registered with the online research company Cross Marketing. Cross Marketing is one of the largest research companies in Japan with 2.95 million active monitors as of January 2022. In the first wave, an online survey was conducted between April 18–19, 2023. In the screening survey, I confirmed participants’ consent and conducted an instructional manipulation check (IMC)^16^ ^17^ to identify inattentive respondents. Participants were instructed to choose “do not start answering” between “start answering” and “do not start answering.” Participants who chose “start answering” were warned to read again carefully and asked to follow the same instruction. I excluded inattentive respondents who did not follow the instructions twice. After the IMCs, participants were asked about their age, gender, place of residence (prefectural level), occupation, marital status, and presence/absence of children. Those who responded with an age not covered by the study, answered “other” for gender, or chose to live outside Japan were excluded from the study.

A total of 2,262 individuals participated in the screening survey. Of these, 224 did not provide consent, 1,279 were excluded because of IMCs, and 11 were not included due to age, gender, or residence. Of the remaining 748, the survey was conducted until 500 valid responses were obtained to match the distribution of gender and age groups in Japan. Short-term respondents were excluded based on the survey company criteria.

The second wave of survey was also conducted until 500 valid responses were obtained. First, between June 6–15, 2023, 491 people who provided valid responses in the first wave of the survey (excluding nine people who were dropped from the survey company’s registry between the two waves) were asked to participate in the second wave of the survey. A total of 459 people participated in the screening survey. Of these, 15 did not provide consent, 129 were excluded due to IMCs, and none were excluded on the basis of age, gender, or residence. Of the remaining 315 respondents, 291 completed the survey.

In addition, new monitors (excluding those identified as inattentive respondents in the first wave of the survey) were recruited between June 13–15, 2023. Of the new monitors, 1,738 participated in the screening; of which 161 did not provide consent, 1,027 were excluded due to IMCs, and eight were excluded on the basis of age, gender, or residence. A total of 209 valid responses were obtained from the remaining 542 participants. Valid respondents who participated in both surveys and those who were new to participate in the second wave of the survey were selected to match the distribution of gender and age groups in Japan. Short-term respondents were also excluded from the second wave in accordance with the survey company’s criteria.

The study finally included 291 participants who completed both the surveys. Survey respondents were awarded points that could be redeemed for products. Those who were excluded due to inattentive responses, age, gender, or residence, and those who stopped responding midway through the main survey parts were awarded points only for the screening part responses.

### Survey Items

The same questionnaire was used in the first and second surveys. I obtained permission from the author of a previous study ^11^ and used the questionnaire items on mask usage and beliefs regarding the associated reasons. The following six items were used as reasons for mask usage in the previous study ^11^:

- Severity: Do you think that your disease condition would be serious if you had COVID-19?
- Protection: Do you think that wearing a mask will keep you from being infected?
- Prevention: Do you think that people who have COVID-19 can avoid infecting others by wearing masks?
- Norm: When you see other people wearing masks, do you think that you should wear a mask?
- Relief: Do you think that you can ease your anxiety by wearing a mask?
- Impulsion: Do you think that you should “do whatever you can” to avoid COVID-19?

It should be noted that norm can be divided into two types: “information effect,” which motivates people to view the behavior of others as correct and behave similarly; and “peer pressure,” which motivates people to behave in accordance with the expectations of others ^18^. Therefore, with permission from the author of the previous study ^11^, the following two questions were also included in this study:

- Information effect: When you see others wearing a mask, do you think it is good behavior to wear a mask?
- Peer pressure: Will you be willing to wear a mask to avoid being blamed by others?

These eight items were rated on a 5-point Likert scale (1 = not at all to 5 = very much).

In the previous study ^11^, participants were enquired about their mask usage since the COVID-19 outbreak (excluding hay fever prevention purposes). However, in this study, they were asked about their mask usage in the past week on a 3-point Likert scale (1 = I have not worn one at all, 2 = I sometimes wear one, and 3 = I usually wear one).

In addition, the participants were also asked about their education and annual household income as individual attributes. Other questions included time of waking and sleeping, records based on day reconstruction method (i.e., time and types of activity events, people they were with, well-being, and mask usage) ^19^, hay fever, items emphasized in diet, smoking habits, and alcohol consumption habits, which were not used in this study. No data were missing. Table 1 lists the individual attributes of the study participants.

**Table 1.**
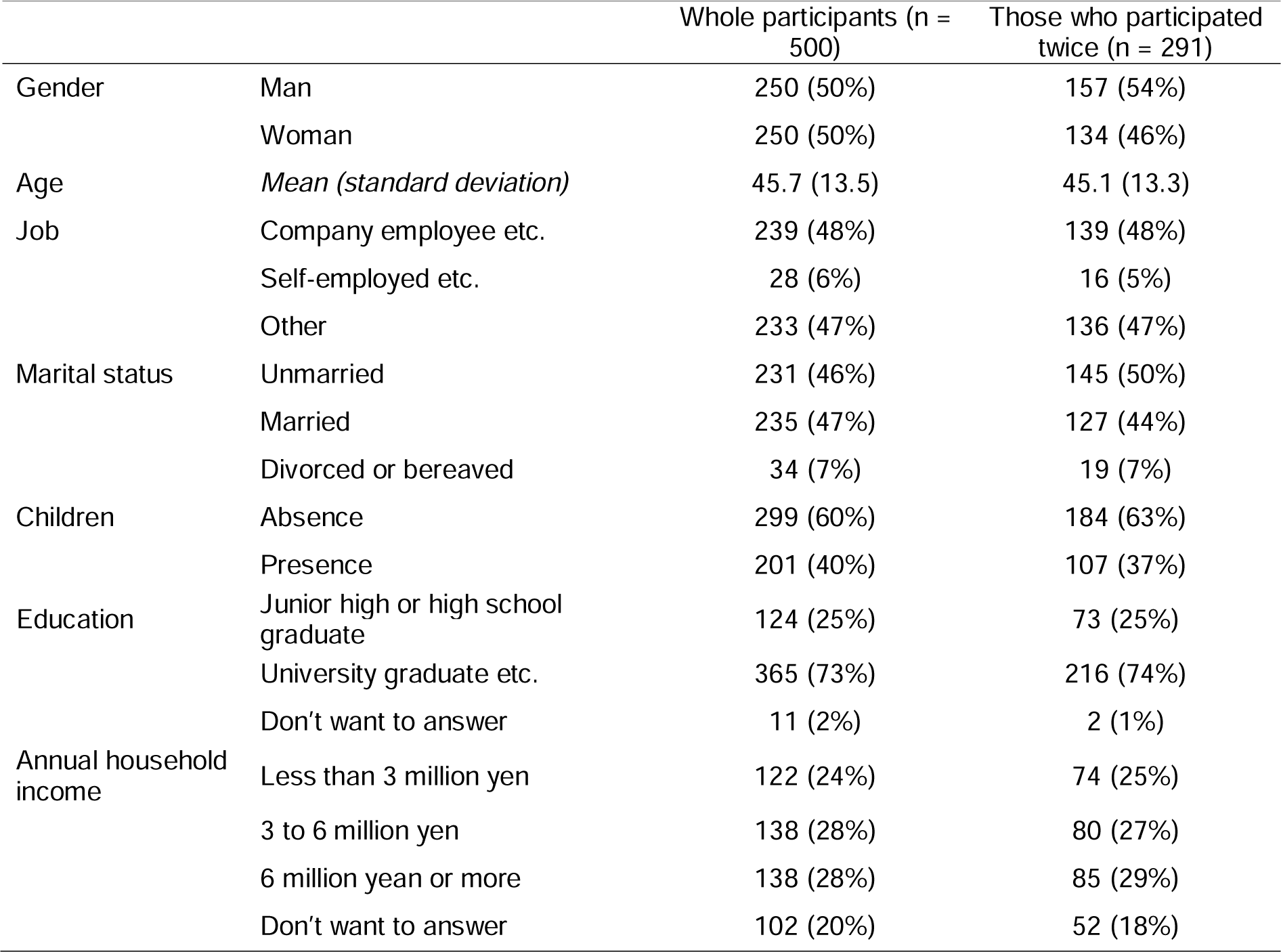
Characteristics of the study participants during the first wave. n: number of participants.

|  |  | Whole participants (n = 500) | Those who participated twice (n = 291) |
| --- | --- | --- | --- |
| Gender | Man | 250 (50%) | 157 (54%) |
|  | Woman | 250 (50%) | 134 (46%) |
| Age | *Mean (standard deviation)* | 45.7 (13.5) | 45.1 (13.3) |
| Job | Company employee etc. | 239 (48%) | 139 (48%) |
|  | Self-employed etc. | 28 (6%) | 16 (5%) |
|  | Other | 233 (47%) | 136 (47%) |
| Marital status | Unmarried | 231 (46%) | 145 (50%) |
|  | Married | 235 (47%) | 127 (44%) |
|  | Divorced or bereaved | 34 (7%) | 19 (7%) |
| Children | Absence | 299 (60%) | 184 (63%) |
|  | Presence | 201 (40%) | 107 (37%) |
| Education | Junior high or high school graduate | 124 (25%) | 73 (25%) |
|  | University graduate etc. | 365 (73%) | 216 (74%) |
|  | Don't want to answer | 11 (2%) | 2 (1%) |
| Annual household income | Less than 3 million yen | 122 (24%) | 74 (25%) |
|  | 3 to 6 million yen | 138 (28%) | 80 (27%) |
|  | 6 million yen or more | 138 (28%) | 85 (29%) |
|  | Don't want to answer | 102 (20%) | 52 (18%) |

### Statistical analysis

Parametric tests were used to ensure consistency with the previous study ^11^. First, t-test was conducted for both first and second waves to investigate the differences in mask usage between the participants in this study (i.e., those who participated in both the first and second waves) and those who participated only in the first or second wave. This was done to check whether the study participants were biased against those who participated only in the first or second wave. A paired t-test was then conducted to analyze the differences in mask usage and beliefs regarding the associated reasons between the first and second waves. The effect sizes for these t-tests were considered small for Cohen’s d of 0.20 and medium for Cohen’s d of 0.50, in accordance with a previous study ^20^.

In addition, a cross-lagged panel model was used to analyze the bi-directional associations between mask usage and beliefs about the associated reasons. First, I considered the initial model of the association between mask usage and beliefs regarding the reasons for mask usage, as shown in Figure 1. The initial model incorporated the information effect and peer pressure instead of norm. Referring to a previous study ^21^, *P* < 0.05 for chi-square, comparative fit index (CFI) ≥ 0.90, root mean square error of approximation (RMSEA) < 0.08, and standardized root mean squared residual (SRMR) < 0.08 were considered acceptable fits for the model. The initial model had a slightly less-than-acceptable fit. Therefore, I changed the model to include the structure of factor analyses regarding beliefs about the associated reasons for mask usage in light of the fact that they were categorized into two types of reasons (i.e., infection risk avoidance and other socio-psychological reasons) in the previous study ^11^ and that correlations were also found between these beliefs in the previous study ^11^ and this study (Table S1). In addition, the model was improved by adding covariates related to individual attributes and removing non-significant paths, except the associations between mask usage and beliefs about the associated reasons between the first and second waves (Figure 2). Bias-corrected 95% confidence intervals (CIs) for path coefficients were calculated using the 1,000 bootstrap method. SPSS and AMOS 28 (IBM, Chicago, IL, U.S.) were used for all the statistical analyses.

**Figure 1.**
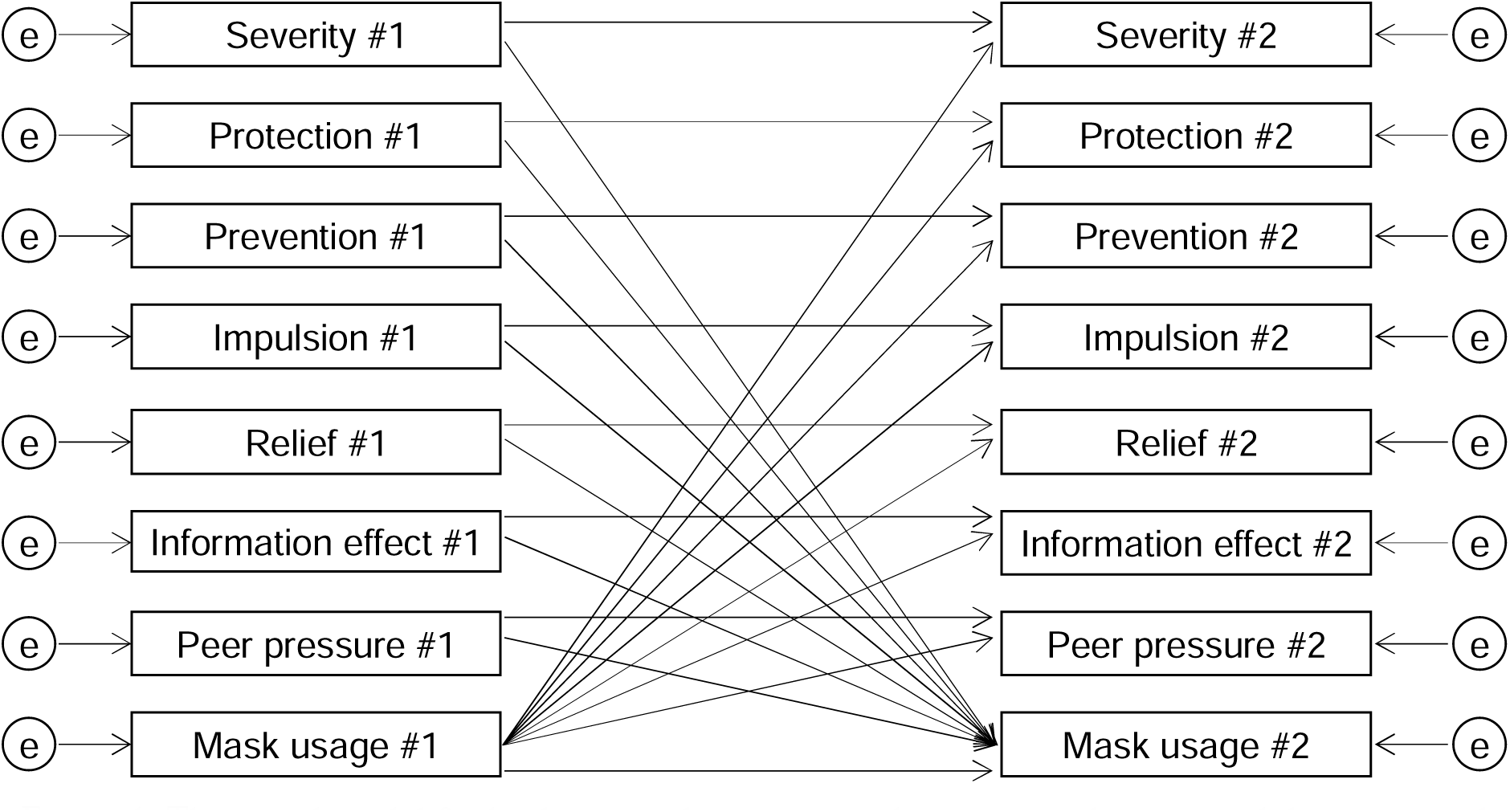
The initial model for bi-directional associations between mask usage and the associated reasons. e: error term, #1: the first wave; #2: the second wave. The paths between the error terms regarding the variables during the first or second wave are omitted in the figure. Chi-square: *P* < 0.001; comparative fit index: 0.96; root mean square error of approximation: 0.09; standardized root mean squared residual: 0.09.

**Figure 2.**
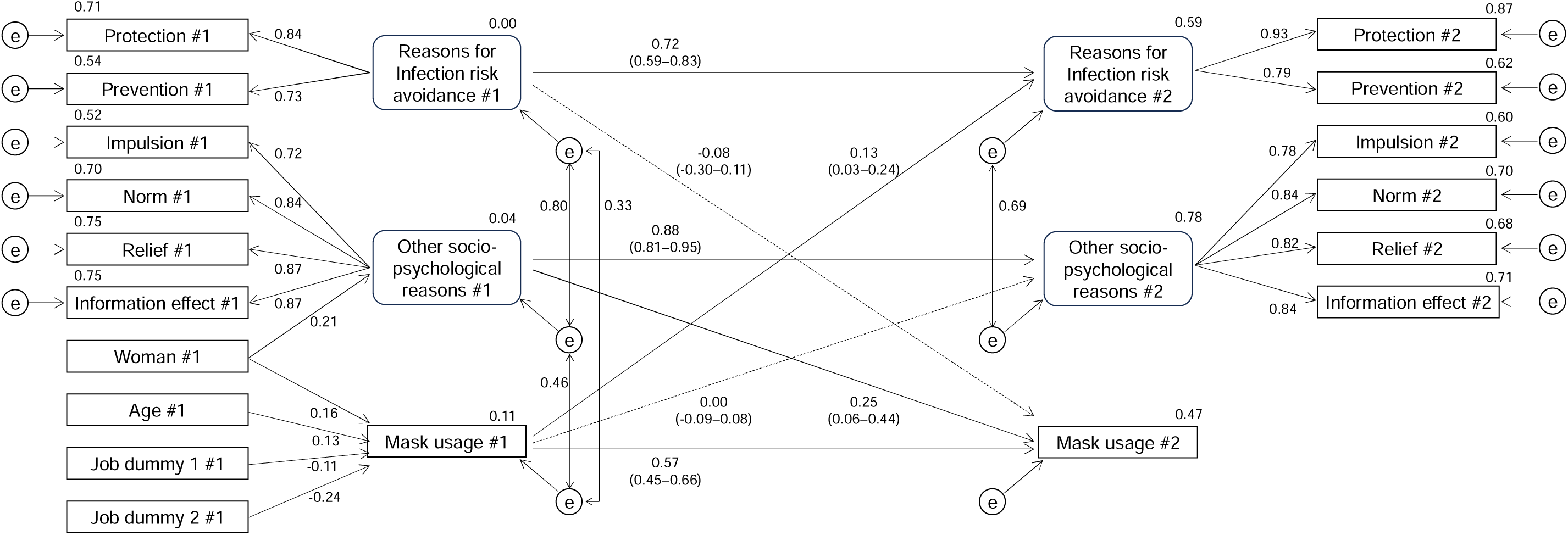
The modified model for bi-directional associations between mask usage and the associated reasons. e: error term, #1: the first wave; #2: the second wave. Job dummy 1 and 2 represents self-employed, etc. and others (ref = company employees etc.). Chi-square: *P* < 0.001; comparative fit index: 0.91; root mean square error of approximation: 0.08; standardized root mean squared residual: 0.05. The values represent standardized estimates. All the paths except dashed lines were significant (P < 0.05). The bias-corrected 95% confidence intervals (CIs) are shown for paths between #1 and #2. The 95% CI is estimated using a bootstrap method of 1000 samples.

## RESULTS

### Changes in mask usage and beliefs about the associated reasons

Mask usage among participants in this study (i.e., those who participated in both the first and second waves) was not significantly different from that among those who participated only in the first or second wave [the first wave: *P* = 0.75, Cohen’s d = −0.03 (95% CI: −0.21–0.15); the second wave: *P* = 0.20, Cohen’s d = 0.12 (−0.06–0.29); Table 2].

**Table 2.** Differences in mask usage between those who participated twice and those who participated once. CI: confidence interval.

|  | Mask usage: mean (standard deviation) |  | Cohen's d (95% CI) | *P* |
| --- | --- | --- | --- | --- |
|  | Those who participated twice (n = 291) | Those who participated once (n = 209) |  |  |
| The first wave | 2.60 (0.61) | 2.62 (0.59) | −0.03 (−0.21–0.15) | 0.75 |
| The second wave | 2.48 (0.69) | 2.40 (0.71) | 0.12 (−0.06–0.29) | 0.20 |

In the first and second waves of the survey, 67% and 59% of the participants reported wearing masks usually, respectively. The arithmetic means (standard deviations) were 2.60 (0.61) and 2.48 (0.69), respectively, indicating a significant decrease (*P* < 0.001, Cohen’s d = −0.23 (−0.35–−0.12) (Table 3). Furthermore, there was a significant decrease in relief and information effects in terms of the beliefs about the associated reasons for mask usage [relief: *P* = 0.046, Cohen’s d = −0.12 (−0.23–0.00); information effect: *P* = 0.018, Cohen’s d = −0.14 (−0.25–−0.02)]. No significant changes were observed in terms of other beliefs.

**Table 3.**
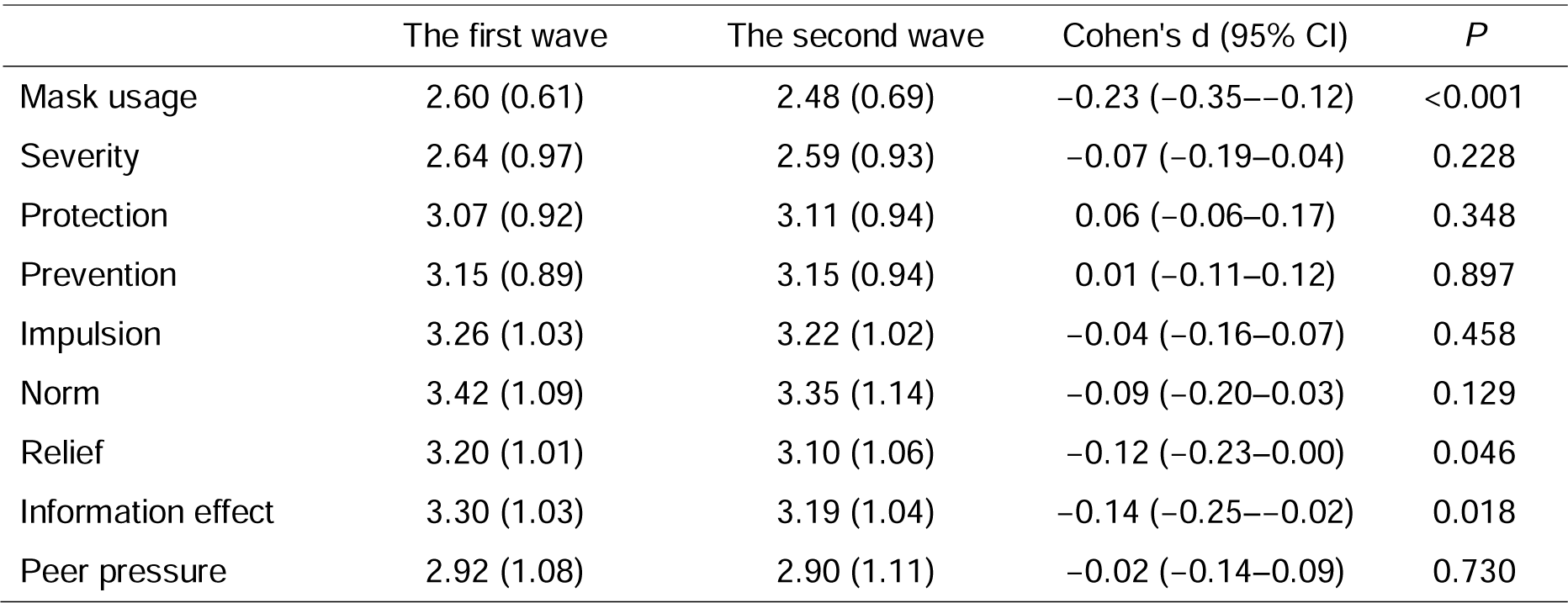
Differences in mask usage and the associated reasons between the first and second waves (n = 291). CI: confidence interval.

### Bi-directional associations between mask usage and beliefs about the associated reasons using a cross lagged panel model

The initial model showed a slightly less-than-acceptable fit, as was revealed by *P* < 0.001 for chi-square, 0.96 for CFI, 0.09 for RMSEA, and 0.09 for SRMR (Figure 1). Considering the strong correlations between beliefs about the associated reasons for mask usage (Table S1), I created an improved model fit, which was determined to be acceptable, with *P* < 0.001 for chi-square, 0.91 for CFI, 0.08 for RMSEA, and 0.05 for SRMR (Figure 2).

In the improved model, the reasons for infection risk avoidance included protection and prevention, whereas other socio-psychological reasons included impulsion, norms, relief, and information effects. The standard estimates for the reasons for infection risk avoidance, other socio-psychological reasons, and mask usage from the first to the second wave were 0.72 (95%CI:0.59– 0.83), 0.88 (0.81–0.95), and 0.57 (0.45–0.66), respectively. Other socio-psychological reasons during the first wave showed a significant path to mask usage during the second wave [standard estimates:0.25 (0.06–0.44)], while the path from reasons for infection risk avoidance during the first wave to mask usage during the second wave was not significant [−0.08 (−0.30–0.11)]. On the other hand, mask usage during the first wave showed a significant path to reasons for infection risk avoidance [0.13 (0.03–0.24)], while the path to other socio-psychological reasons during the second wave was not significant [0.00 (−0.09–0.08)].

## DISCUSSION

In this study, I conducted two-wave surveys focusing on the bi-directional associations between mask usage and beliefs about the associated reasons before and after the downgrading of the legal status of COVID-19 in Japan.

Mask usage showed no significant differences in the first and second waves between the participants in this study (i.e., those who participated in both the first and second waves) and those who participated once in either of the surveys. The effect sizes were also judged to be small. Therefore, the participants in this study were not considered to be biased in comparison to the overall respondents targeted in the surveys.

Mask usage significantly decreased from the first to the second wave, but the effect size was small. A total of 67% and 59% of the participants reported wearing masks usually before (i.e., April 18–19, 2023) and after the downgrading of the legal status of COVID-19 (i.e., June 6–15, 2023), respectively, indicating that mask usage proportion was still high in Japan. This result was slightly higher than the 51% reported in a previous study conducted in Japan between March 26–31, 2020 ^11^. Another survey ^8^ showed that the proportions of Japanese respondents who reported “wearing a mask in public places within the past two weeks” were 86% (September 2022), 71% (March 15–22, 2023), 68% (April 12–19), and 66% (May 10–17), and the results of this study were similar to those obtained in the previous survey. The results of the first wave of surveys conducted in this study were similar to those of the first wave of surveys conducted by the government. Although the first wave of surveys in this study was conducted between the government’s policy change that mask wearing was based on individual judgment (i.e., March 13, 2023) and the downgrading of the legal status of COVID-19 (May 8), some of the mask wearers, as of September 2022, had already stopped using masks during the time of the first wave of this study. Among beliefs about the reasons for mask usage, relief and information effects also declined significantly; however, the effect sizes were small. The cross-lagged panel model also showed high standard estimates for reasons of mask usage from the first to the second wave, representing small intra-individual variations. This revealed that the impact of downgrading the legal status of COVID-19 on mask usage and the reasons for this were limited.

The cross-lagged panel model showed that socio-psychological reasons other than infection risk avoidance, in the first wave were significantly associated with mask usage in the second wave. This indicates that those who perceived socio-psychological factors, such as impulse, norms, relief, and information effect, as less important reasons for mask usage in the first wave were significantly less likely to wear masks in the second wave. The results of this study are consistent with previous findings ^11^ ^12^ that norm and relief influences mask wearing in Japan. However, while mask usage was reported to be a social norm in Hong Kong during the H1N1 influenza outbreak ^22^, and mask usage in Switzerland was strongly associated with infection risk avoidance after the COVID-19 outbreak ^13^, the findings of this study may not be applicable to other countries, such as Western countries. Using ethnographic interviews and participant-observations at public sites in Japan, the United States, and China ^23^, social pressure in Japan, state pressure in China, and politically based individual choices in the United States were identified as important factors in mask wearing.

In contrast, mask usage in the first wave was significantly associated with the reasons for infection risk avoidance and constituting protection and prevention in the second wave. This indicates that mask users reinforced the belief that masks can avert infection risk or that non-mask users came to believe that masks are ineffective in avoiding the infection risk. The mechanism by which wearing a mask influences one’s beliefs about the associated reason can be explained using the self-perception theory ^14^, which states that one’s perception of oneself wearing (or not wearing) a mask leads to one’s belief that avoiding infection risk is important (or not important) as a straightforward interpretation of one’s behavior. It is worth noting that while there was a significant association between mask usage and the reasons for infection risk avoidance, the association of mask usage with other socio-psychological reasons was not significant. One possible interpretation for this is that individuals apply their own beliefs to the importance of infection risk avoidance, which seems to be a more logical and rational reason.

This study has implications for advancing mask-usage strategies. A previous study ^11^ showed that people’s sense of norm and relief could be the driving forces behind the promotion of mask usage in Japan, and highlighted the importance of using nudge messages based on social motivations and presenting risk information in accordance with subjective emotions. This study also showed the significance of risk communication based on socio-psychological reasons other than infection risk avoidance, such as norms and relief, even in situations where mask usage proportion declined. Furthermore, this study also showed that unmasking led to a decrease in the rationale for infection risk avoidance. This means that, as mask usage continues to decline, awareness of the importance of infection risk avoidance through masks may decline further. Risk communication strategies that focus on the importance of infection risk avoidance through masks will be challenging when promoting mask-wearing during another wave of COVID-19 and outbreaks of emerging infectious diseases. Nevertheless, I do not intend to imply that widespread dissemination of information regarding the fact that wearing masks can avert the risk of infection is pointless. The dissemination and sharing of information regarding the central role of mask wearing in infection risk avoidance may support socio-psychological reasons such as norms and information effects in the public. Regardless of whether mask-wearing is promoted or mitigated, risk communication should be considered in accordance with the fact that risk-coping behaviors involve a combination of infection risk avoidance and other socio-psychological reasons.

This study had a few limitations. First, this study used online surveys, which may have biased the participants of this study. However, the participants were given points after the surveys, which had the advantage of providing incentives for participants who were not interested in the topics of this study to participate in the survey. In addition, the cross-lagged panel model was adjusted to incorporate covariates such as age, gender, and occupation. Second, the findings of this study were based in Japan, and caution should be exercised when applying these findings to other countries. As already discussed, the association between mask usage and beliefs about the associated reasons may differ in other countries. Third, although the longitudinal study design and cross-lagged panel model discussed the direction of the associations between mask usage and beliefs about the associated reasons, they did not lead to the identification of a causal relationship. Future randomized controlled trials in multiple countries are expected to deepen the findings regarding the associations between mask usage and beliefs about the associated reasons, and to implement risk communication tailored to the target country and infection situation.

Despite these limitations, based on two waves of surveys conducted before and after the downgrading of the legal status of COVID-19 in Japan, this study found that there were bi- directional associations between mask usage and beliefs about the associated reasons. Moreover, different directions of associations of mask usage with infection risk avoidance and other socio- psychological reasons were also found.

## Data Availability

All data produced in the present study are available upon reasonable request to the authors.

## Acknowledgements

We would like to thank Editage (www.editage.com) for English language editing. We are also grateful for helpful discussions: Dr. Tomoyuki Kobayashi (Fukushima Medical University), Dr. Mao Yagihashi (Osaka University), Ms. Misato Okaneya (Osaka University), Dr. Mei Yamagata (Doshisha University), Dr. Asako Miura (Osaka University), and Dr. Kazuya Nakayachi (Doshisha University).

