## Supplemental Table S1 for "Bi-directional associations between mask usage and the associated reasons before and after the downgrading of the legal status of COVID-19 in Japan: A longitudinal study"

Table S1. Correlation matrix (Pearson' s *R*) among mask usage and the associated reasons (n = 291). Upper right: the first wave; Lower left: the second wave.

| | Mask usage | Severity | Protection | Prevention | Impulsion | Norm | Relief | Information effect | Peer pressure |
|---|---|---|---|---|---|---|---|---|---|
| Mask usage | - | 0.186** | 0.257** | 0.230** | 0.375** | 0.382** | 0.396** | 0.361** | 0.264** |
| Severity | 0.104 | - | 0.277** | 0.145* | 0.325** | 0.344** | 0.236** | 0.314** | 0.250** |
| Protection | 0.257** | 0.345** | - | 0.628** | 0.563** | 0.513** | 0.549** | 0.535** | 0.244** |
| Prevention | 0.221** | 0.187** | 0.737** | - | 0.468** | 0.427** | 0.522** | 0.478** | 0.253** |
| Impulsion | 0.310** | 0.330** | 0.590** | 0.501** | - | 0.590** | 0.602** | 0.618** | 0.189** |
| Norm | 0.348** | 0.352** | 0.569** | 0.471** | 0.667** | - | 0.725** | 0.744** | 0.411** |
| Relief | 0.273** | 0.291** | 0.602** | 0.542** | 0.599** | 0.682** | - | 0.752** | 0.291** |
| Information effect | 0.300** | 0.450** | 0.577** | 0.494** | 0.649** | 0.713** | 0.710** | - | 0.344** |
| Peer pressure | 0.206** | 0.370** | 0.381** | 0.325** | 0.391** | 0.586** | 0.457** | 0.510** | - |

* $P < 0.05$; ** $P < 0.01$.
